## Supplementary Appendices for "Let’s just ask them. Perspectives on urban dwelling and air quality: a cross-sectional survey of 3,222 children, young people and parents": Supplementary Material.pdf

Supplementary Appendix 1: Most successful advertisement for each target city

| **Milan**<br>*Most successful creative* | **Mexico City**<br>*Most successful creative* | **Los Angeles**<br>*Most successful creative* | **London**<br>*Most successful creative* |
|---|---|---|---|
| 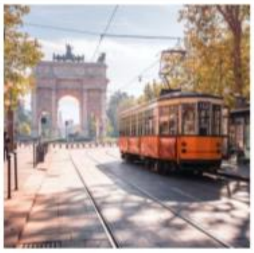 | 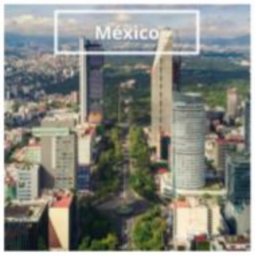  | 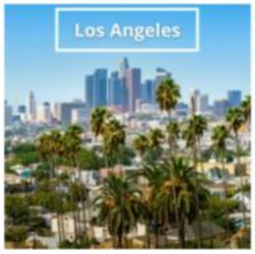  | 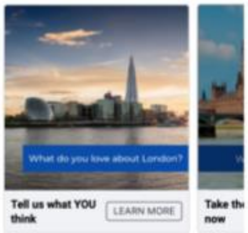  |
| 0.95% CTR | 1.40% CTR | 3.97% CTR | 1.43% CTR (carousel) |

| **Freetown**<br>*Most successful creative* | **Dhaka**<br>*Most successful creative* | **Dar es Salaam**<br>*Most successful creative* | **Bhubaneswar**<br>*Most successful creative* |
|---|---|---|---|
| 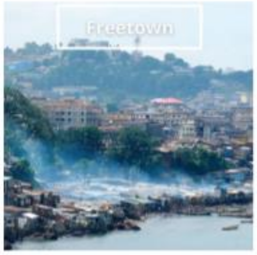  | 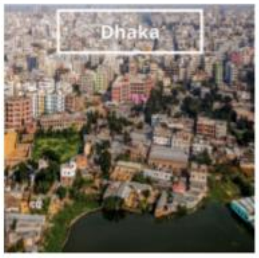  | 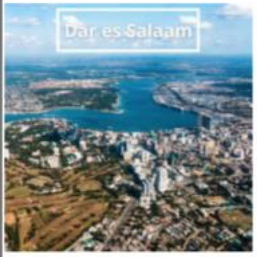  | 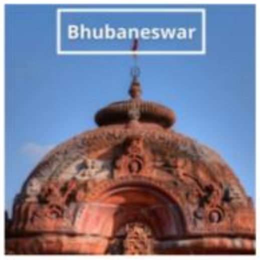  |
| 1.63% CTR | 1.33% CTR | 1.98% CTR | 0.90% CTR |

| Lahore<br>*Most successful creative* | Jaipur<br>*Most successful creative* | Harare<br>*Most successful creative* | Glasgow<br>*Most successful creative* |
|---|---|---|---|
| 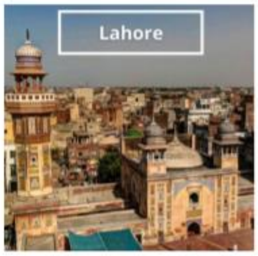 | 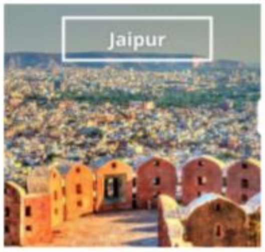 | 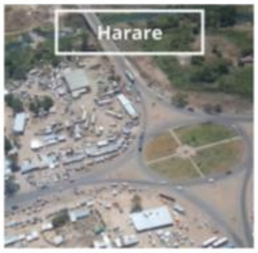 | 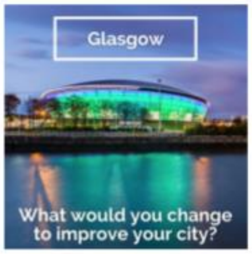 |
| 1.69% CTR | 1.93% CTR | 1.65% CTR | 1.69% CTR (carousel) |

| Tamale<br>*Most successful creative* | Quito<br>*Most successful creative* | Quezon<br>*Most successful creative* | Nairobi<br>*Most successful creative* |
|---|---|---|---|
| 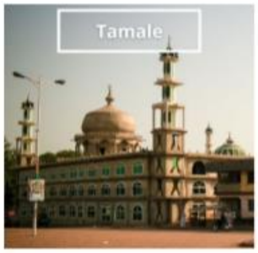 | 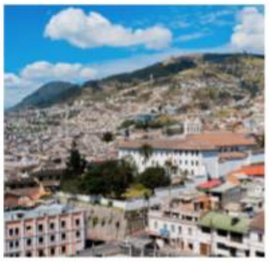 | 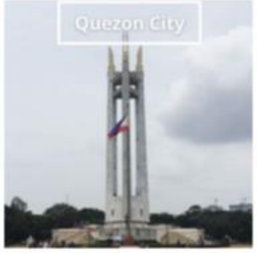 | 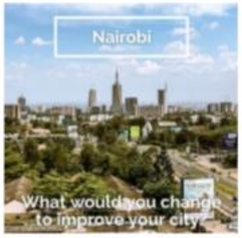 |
| 2.05% CTR | 1.01% CTR | 0.51% CTR | 1.61% CTR |

Supplementary Appendix 2: Detailed methods for population recruitment, pretesting methods, and data handling Population Recruitment

Recruitment advertisements were targeted at people living in 16 cities in the respective languages: London (English), Glasgow (English), Milan (Italian), Quezon City (English), Los Angeles (English), Nairobi (Swahili/Sheng), Quito (Spanish), Jaipur (Hindi and Urdu), Bhubaneswar (Hindi), Dar es Salaam (Swahili/Sheng), Tamale (English), Lahore (Urdu), Dhaka (Bangla), Free Town (English), Mexico City (Spanish), Harare (English). These cities were not randomly selected, rather chosen due to existing connection to the CCC project (London, Nairobi, and Harare) or as the hosts of the 2021 COP26 Summit (Glasgow and Milan). The additional cities were chosen to reflect a variety of global settings, population sizes, and levels of development in both the Global North and South. All 16 cities were used in the co-benefits analysis completed by the CCC team, to quantify the effects of reducing urban AP on child health.

Respondents were recruited to the survey through promoted social media advertisements that targeted young people or parents in the 16 target locations, based on location and demographic data held by the social media platforms (22). Recruitment advertisements were co-developed with a specialist social media marketing agency (23), and were designed to appeal to a wide variety of young people (not just those already concerned about urban/environmental health) using a variety of short messages and images (Supplementary Supplementary Appendices

Supplementary Appendix *1*). The advertisements were pre-tested to optimise engagement for each target group and location based on clicks (rather than survey completes), to reduce the risk of introducing additional responder bias beyond that already inherent due to variable completion rates. Several adjustments were made throughout the campaign to attempt to limit bias in the sample. To identify more potential respondents from less represented areas, location radius was increased for cities with a higher cost per click and adverts which produced the lowest click-through rates were refreshed with new imagery. To reduce over representation in Tamale, Harare, and Dhaka advertisements were paused in these locations on October 5$^{th}$, 2021 as they had produced the largest volume of eligible and consenting responses at that time. Informed consent, developed with reference to guidelines in *The Global Kids Online Research Toolkit* (24), was sought from all respondents in a relevant language before they were included into the final sample (web appendix 4).

**Pretesting Methods**

The English-language survey instrument was pre-tested in two phases to evaluate its reliability and validity and ensure data quality. Within each phase of pre-testing, feedback on the survey instrument was collected using a Google Form (25), in parallel time with the completion of the survey instrument. Feedback was integrated into the draft Typeform survey instrument after each phase of pre-testing. The survey was translated into Spanish, French, Swahili/Sheng, Hindi, Urdu, Italian, Bangla, Arabic, and Chinese through a combination of an initial automated translation (using Google Translate (26)) followed by proof reading and correction by native language speakers in each language. Data were collected from August 17$^{th}$ to October 10$^{th}$ 2021 on respondents' own devices.

**Data Storage**

All responses were stored on the Typeform.com servers (hosted in a Virtual Private Cloud by Amazon Web Services) and processed and analysed on password-protected encrypted devices. All data collection tools, anonymised raw data, and data analysis code are available on the LSHTM Data Compass Site (14) mirrored on Figshare (27). The dataset has been assigned a unique DataCite DOI

(28). Confidentiality of survey responses was maintained throughout; no personally identifiable data was collected. Age in years was collected, and the location was recorded at the city level. In addition, no IP address or other locator/identifiers (like cookies) were captured.

Supplementary Appendix 3: Survey instrument

| *Description or Typeform survey text* | *Answer options* |
|---|---|
| *"How old are you?"* | ***Open text*** |
| *"Are you or your partner currently pregnant?"* | ***Single choice:***<br>*Pregnant*<br>*Not pregnant* |
| *"Where do you live?"* | ***Single choice:***<br>*Bubaneswar (India)*<br>*Dar es Salaam (Tanzania)*<br>*Dhaka (Bangladesh)*<br>*Freetown (Sierra Leone)*<br>*Glasgow (Scotland)*<br>*Jaipur (India)*<br>*Lahore (Pakistan)*<br>*London (England)*<br>*Los Angeles (USA)*<br>*Milan (Italy)*<br>*Quito (Ecuador)*<br>*Tamale (Ghana)* |
| *"What are the BEST things about living in your city/town?"* | ***Multiple selection, up to three:***<br>*The people*<br>*Places to play*<br>*Being close to school or work*<br>*There are many things to do*<br>*The shops and restaurants*<br>*It's easy to get around*<br>*Healthcare*<br>*Being close to my family*<br>*The climate and environment*<br>*Work opportunities for my family*<br>*Access to green space, like parks*<br>*Other* |
| *"Now, what are the WORST things about living in your city/town?"* | ***Multiple selection, up to three:***<br>*It doesn't always feel safe*<br>*The people; it's not friendly*<br>*The traffic/congestion*<br>*The noise*<br>*Not enough places to play*<br>*Not enough places to meet friends*<br>*Shortage of work opportunities for my family*<br>*It's hard to get around*<br>*Not easy enough to get healthcare*<br>*The pollution* |

| | |
|---|---|
| | *It's too crowded*<br>*Not enough green space like parks*<br>*Other* |
| *"Do you have any comments to add (about your city in general)?"* | ***Open text*** |
| *"We are interested in what you think about the air quality where you live"* | ***Single choice:***<br>*Numeric 0-10* |
| *"What do you think are the main sources of AP where you live"* | ***Multiple selection, unlimited:***<br>*Factories*<br>*Burning of rubbish*<br>*Motor transport (cars, buses, lorries)*<br>*Construction/building work*<br>*Pollution blown into the city (from outside)*<br>*Household cooking (cooking fires/stoves)*<br>*Household heating (boilers, wood fires etc)*<br>*Agriculture/farming*<br>*Other* |
| *"Do you have any comments to add (about the air quality in your city)?"* | ***Open text*** |
| *"In general, is your city becoming a nicer place to live, or a less nice place to live?"* | ***Single choice:***<br>*Becoming a nicer place to live*<br>*Staying the same*<br>*Becoming a worse place to live* |
| *"Why do you say that?"*<br>*Qualitative responses to the nicer_city question* | ***Open text*** |
| *"If you were the mayor of your town/city, how would you…Improve the city generally, especially for children and young people?"* | ***Open text*** |

| | |
|---|---|
| *"And, how would you improve air quality where you live?"* | ***Open text*** |
| *"Do you have any more comments about any of these issues?"* | ***Open text*** |

Supplementary Appendix 4: Language, consent, and eligibility questions

| *Variable Name* | *Description or Typeform survey text* | *Answer options* |
|---|---|---|
| *Lang* | *Hello presented in each language* | ***Single Choice:***<br><br>*Hello*<br>*Hola*<br>*Bonjour*<br>*Niaje*<br>*नमस्ते*<br>*ہیلو*<br>*Ciao*<br>*হ্যালো*<br>*أهلا*<br>*你好* |
| *Eligibility 1* | *Before we start, we need to tell you about this research so you can decide if you want to take part.*<br><br>***Who are we?*** *We are a group of researchers from London School of Hygiene and Tropical Medicine who are asking children, young people and parents from all over the world to tell us what they think of their cities. We think children's voices and opinions are not included enough, and want to try to address this.*<br><br>***Why are you seeing this survey?*** *We paid for posts to appear in Facebook/Instagram feeds of people like you who live in your city. (This is based on the information that Facebook/Instagram holds about you, not any information we have – we do not have, or have access to, any of your personal information at all).*<br><br>***How long is it?*** *We have 10 questions for you, that will take you less than 5 minutes to answer.*<br><br>***How can I contact you if I have questions?*** *You can email us (the study leads Prof Alan Dangour and Dr Rob Hughes) at if you have any questions about this survey.*<br><br>***To take part you must be:***<br><br>*Aged between 13-25 years old AND live in a town/city, OR*<br><br>*A parent of someone under 13 years old AND live in a town/city, OR*<br><br>*Aged over 18, you or your partner are expecting a baby AND live in a town/city*<br><br>***Select the option that best describes you:*** | ***Single choice:***<br><br>*Aged between 13-25 years old*<br><br>*A parent of someone under 13 years old*<br><br>*Aged over 18, you or your partner are expecting a baby*<br><br>*None of these describe me* |

| | | |
|---|---|---|
| *Consent 1* | *Participation is completely voluntary; you don't have to take part and you can stop at any time* | ***Single choice:***<br><br>*OK, I want to complete the survey*<br><br>*No, I don't want to complete the survey* |
| *Consent 2* | *Your answers are anonymous – that means nobody will know who you are and nobody will be able to connect your answers back to you (including the researchers)* | ***Single choice:***<br><br>*OK, I want to complete the survey*<br><br>*No, I don't want to complete the survey* |
| *Consent 3* | *We will write research reports, blogs, and social media posts based on what we learn from all survey responses combined together* | ***Single choice:***<br><br>*OK, I want to complete the survey*<br><br>*No, I don't want to complete the survey* |
| *Consent 4* | *Storing and sharing data: We will keep your answers securely on our computers, and we might also share them with other scientists in the future too (we will never have or share any information that could identify you individually)* | ***Single choice:***<br><br>*OK, I want to complete the survey*<br><br>*No, I don't want to complete the survey* |
| *Consent 5* | *You will not get any reward or prize for taking part*<br><br>*(To show our thanks, you will receive a certificate of participation at the end that you can print out.)* | ***Single choice:***<br><br>*OK, I want to complete the survey*<br><br>*No, I don't want to complete the survey* |
| *Question 1* | *Do you have any questions?* | ***Single choice:***<br><br>*Yes - Take me to the FAQs*<br><br>*No - START survey* |
| *FAQs* | *FAQs*<br><br>***Why are you doing this work?***<br><br>*We are really keen to learn what children, young people and parents (and people who are soon to become parents) think of their city. We think children's voices and opinions are not included enough, and want to try to address this.* | ***Single choice:***<br><br>*Yes - get in touch*<br><br>*No – START survey* |

| | |
|---|---|
| | ***2. How did you select me?***<br><br>*We want to hear from people who live in your city who are children/young people, parents of children or expecting a baby soon. We paid for posts to appear in Facebook/Instagram feeds of people like you who live in your city. (This is based on the information that Facebook/Instagram holds about you, not any information we have – we do not have, or have access to, any personal information at all).*<br><br>***3. Why do I not get anything in return for completing the survey?***<br><br>*Firstly, this is a very short survey – it will probably only take you up to 6-7 minutes to complete it. Also, sending you a thank you for your time would require collecting personal information about you, like your phone number or email address; we don't want to collect and store that information, so thought it would be simpler to simply THANK YOU for your time if you do decide to complete the survey. (There is a certificate you can print at the end if you'd like to).*<br><br>***4. Who are you?***<br><br>*We are researchers at the London School of Hygiene and Tropical Medicine (lead researchers Prof Alan Dangour and Dr Rob Hughes), in the United Kingdom. This work is funded by the Botnar Foundation (which is based in Switzerland).*<br><br>***5. How will you keep my answers private?***<br><br>*Firstly, we are deliberately not collecting any personal information which could identify you from the many thousands of responses we're hoping to collect. Secondly, all the responses are kept safely on a highly secure computer.* |

Supplementary Appendix 5: Survey completions by country

| Country | Frequency | Percent |
|---|---|---|
| Bangladesh | 866 | 26.88 |
| India | 337 | 10.46 |
| Pakistan | 325 | 10.09 |
| Ghana | 301 | 9.34 |
| Ecuador | 288 | 8.94 |
| Zimbabwe | 276 | 8.57 |
| Kenya | 174 | 5.4 |
| United Kingdom | 170 | 5.28 |
| Sierra Leone | 160 | 4.97 |
| Tanzania | 96 | 2.98 |
| Mexico | 90 | 2.79 |
| Italy | 71 | 2.2 |
| Philippines | 57 | 1.77 |
| United States of America | 11 | 0.34 |
| Total | 3,222 | 100 |

Supplementary Appendix 6: Respondents by self-reported town

| Town | Frequency | Percent | Town | Frequency | Percent |
|---|---|---|---|---|---|
| Dhaka † | 858 | 26.63 | Chitral | 1 | 0.03 |
| Lahore † | 315 | 9.78 | Chitungwiza | 1 | 0.03 |
| Tamale † | 300 | 9.31 | Cobham | 1 | 0.03 |
| Quito † | 288 | 8.94 | Cover | 1 | 0.03 |
| Harare † | 271 | 8.41 | Echague | 1 | 0.03 |
| Jaipur † | 225 | 6.98 | Entebbe** | 1 | 0.03 |
| Nairobi † | 169 | 5.25 | Firozabad | 1 | 0.03 |
| Freetown † | 159 | 4.93 | Groningen | 1 | 0.03 |
| Glasgow † | 113 | 3.51 | Haroonabad | 1 | 0.03 |
| Bhubaneswar † | 104 | 3.23 | Jhang | 1 | 0.03 |
| Dar es Salaam † | 96 | 2.98 | Kajiado | 1 | 0.03 |
| Mexico City † | 89 | 2.76 | Khulna | 1 | 0.03 |
| Milan † | 69 | 2.14 | Kuchaman | 1 | 0.03 |
| London † | 56 | 1.74 | Mardan | 1 | 0.03 |
| Quezon City † | 36 | 1.12 | Marikina | 1 | 0.03 |
| Manila | 13 | 0.4 | Masvingo | 1 | 0.03 |
| Los Angeles † | 11 | 0.34 | Monza | 1 | 0.03 |
| missing* | 4 | 0.12 | Muranga | 1 | 0.03 |
| Narayanganj | 3 | 0.09 | Mymensingh | 1 | 0.03 |
| Churu | 2 | 0.06 | Narowal | 1 | 0.03 |
| Sialkot | 2 | 0.06 | New York | 1 | 0.03 |
| Victoria Falls | 2 | 0.06 | Okara | 1 | 0.03 |
| Alwar | 1 | 0.03 | Rourkela | 1 | 0.03 |
| Antipolo | 1 | 0.03 | San Jose Del Monte | 1 | 0.03 |
| Atrai | 1 | 0.03 | San Juan | 1 | 0.03 |
| Bahawalnagar | 1 | 0.03 | Sargodha | 1 | 0.03 |
| Baripada | 1 | 0.03 | Sirsa | 1 | 0.03 |
| Bomet | 1 | 0.03 | Valenzuela | 1 | 0.03 |
| Busia | 1 | 0.03 | Yeji | 1 | 0.03 |
| Caloocan | 1 | 0.03 | **Total** | **3,222** | **100** |

*people who did not explicitly state a town and could not impute from paid media advert campaign meta data

**individual reported living in Entebbe Uganda, but paid media advert campaign meta data identified them as living in Nairobi Kenya. The response was clustered with Nairobi responses.

† City included in the social media campaign advertisements

Supplementary Appendix 7: Distribution of respondents (n=3,222) and focal cities (n=16) by pm2.5 and income indicators

| $PM_{2.5}$ quartile and represented cities | n(%) of n=3,222 sample | Mean (sd) pm2.5 (ug/m3) | World Bank income group | Gross domestic product per capita (USD) |
|---|---|---|---|---|
| **Quartile 1** | **113 (4%)** | **8.00 (0)** | | |
| Glasgow | 113 | 8.00 | High | 34,358 |
| **Quartile 2** | **68 (2%)** | **12.00 (0)** | | |
| London | 57 | 12.00 | High | 58,827 |
| Los Angeles | 11 | 12.00 | High | 57,577 |
| **Quartile 3** | **1,045 (32%)** | **19.86 (2.21)** | | |
| Quito | 288 | 18.0 | Upper-middle | 6,368 |
| Nairobi | 174 | 17.00 | Lower-middle | 6,344 |
| Quezon City | 57 | 18.00 | Lower-middle | 8,482 |
| Freetown | 160 | 21.63 | Low | 1,079 |
| Mexico City | 90 | 22.00 | Upper-middle | 22,587 |
| Harare | 276 | 22.25 | Lower-middle | 1,386 |
| **Quartile 4** | **1,996 (62%)** | **63.03 (18.88)** | | |
| Milan | 71 | 27.00 | High | 51,768 |
| Dar es Salaam | 96 | 29.08 | Lower-middle | 5,699 |
| Tamale | 301 | 55.00 | Lower-middle | 5,150 |
| Dhaka | 866 | 57.00 | Lower-middle | 7,712 |
| Lahore | 325 | 68.00 | Lower-middle | 3,144 |
| Bhubaneswar | 106 | 83.09 | Lower-middle | 1,300 |
| Jaipur | 231 | 105.00 | Lower-middle | 1,500 |

Supplementary Appendix 8: Demographics for full sample and each subpopulation

| | Total<br>n (%) | Age<br>Mean (SD) | Parent of a Child<br>Aged <13<br>n(%) | Young People<br>Aged 13-25<br>n (%) | Expectant Parents<br>n (%) |
|---|---|---|---|---|---|
| 'Best' | 2,939 (100%) | 24 (8.26) | 496 (17%) | 2,187 (74%) | 267 (9%) |
| 'Worst' | 3,051 (100%) | 24 (8.50) | 508 (17%) | 2,266 (74%) | 277 (9%) |
| 'Pollution' | 3,056 (100%) | 24 (8.50) | 509 (17%) | 2,270 (74%) | 277 (9%) |
| 'Air Quality' | 3,054 (100%) | 24 (8.50) | 509 (17%) | 2,267 (74%) | 278 (9%) |
| 'Nicer City' | 3,061 (100%) | 24 (8.49) | 509 (17%) | 2,273 (74%) | 279 (9%) |
| **Full Sample** | **3,222 (100%)** | **24 (8.26)** | **509 (16%)** | **2,430 (75%)** | **283 (9%)** |

Supplementary Appendix 10: Percentage of total n respondents that reported each item within the top 3 'best' aspects of their cities, stratified by $PM_{2.5}$ quartile, age bucket, and respondent group

| | Total n (100%*) | Being close to my family | There are many things to do | Being close to school or work | The shops and restaurants | Work opportunities for my family | The People | It's easy to get around | The climate and environment | Access to green spaces, like parks | Healthcare | Places to Play | Other |
|---|---|---|---|---|---|---|---|---|---|---|---|---|---|
| **Full sample** | 2,939 (100%) | 30% | 27% | 27% | 24% | 22% | 22% | 21% | 18% | 14% | 13% | 4% | 3% |
| **$PM_{2.5}$ Quartile** | | | | | | | | | | | | | |
| 1 | 109 (100%) | 21% | 33% | 39% | 45% | 10% | 36% | 25% | 6% | 33% | 20% | 4% | 4% |
| 2 | 64 (100%) | 13% | 45% | 25% | 39% | 20% | 16% | 30% | 6% | 23% | 17% | 3% | 0% |
| 3 | 863 (100%) | 32% | 24% | 22% | 16% | 19% | 24% | 17% | 32% | 14% | 9% | 3% | 2% |
| 4 | 1,903 (100%) | 29% | 27% | 28% | 26% | 24% | 20% | 22% | 13% | 12% | 14% | 4% | 1% |
| **Age bucket** | | | | | | | | | | | | | |
| Unknown | 9 (100%) | 44% | 11% | 0% | 22% | 0% | 44% | 0% | 33% | 11% | 0% | 11% | 0% |
| 13-16 | 303 (100%) | 24% | 30% | 28% | 33% | 17% | 19% | 18% | 16% | 19% | 14% | 5% | 2% |
| 17-19 | 603 (100%) | 28% | 29% | 31% | 27% | 19% | 20% | 18% | 16% | 12% | 13% | 5% | 1% |
| 20-25 | 1,411 (100%) | 28% | 27% | 25% | 23% | 22% | 22% | 23% | 18% | 14% | 12% | 3% | 1% |
| 25+ | 613 (100%) | 37% | 23% | 25% | 21% | 28% | 23% | 19% | 19% | 14% | 15% | 2% | 1% |
| **Respondent group** | | | | | | | | | | | | | |
| Parent or expectant | 762 (100%) | 34% | 23% | 25% | 20% | 27% | 21% | 19% | 19% | 15% | 14% | 4% | 1% |
| Young person | 2,177 (100%) | 28% | 28% | 27% | 25% | 21% | 22% | 21% | 17% | 14% | 13% | 4% | 1% |

*Stratified percentages add up to over 100%

Supplementary Appendix 11: The percentage of total n respondents that reported each item within the top 3 'worst' aspects of their cities, stratified by $PM_{2.5}$ quartile, age bucket, and respondent group

| | Total n (100%*) | The traffic/ congestion | The pollution | Shortage of work opportunities for my family | It's too crowded | It doesn't always feel safe | The noise | Not enough green spaces like parks | Not easy enough to get healthcare | Not enough places to play | It's hard to get around | Not enough places to meet friends | The people; it's not friendly | Other |
|---|---|---|---|---|---|---|---|---|---|---|---|---|---|---|
| **Full sample** | 3,051 (100%) | 49% | 39% | 23% | 23% | 20% | 16% | 14% | 14% | 11% | 7% | 5% | 5% | 5% |
| **$PM_{2.5}$ Quartile** | | | | | | | | | | | | | | |
| 1 | 113 (100%) | 41% | 34% | 12% | 7% | 54% | 14% | 7% | 4% | 7% | 20% | 14% | 10% | 10% |
| 2 | 67 (100%) | 48% | 42% | 1% | 33% | 45% | 16% | 9% | 4% | 4% | 7% | 7% | 9% | 3% |
| 3 | 898 (100%) | 51% | 31% | 36% | 20% | 27% | 11% | 11% | 23% | 4% | 7% | 5% | 4% | 5% |
| 4 | 1973 (100%) | 48% | 42% | 18% | 24% | 14% | 18% | 16% | 10% | 14% | 6% | 4% | 5% | 4% |
| **Age bucket** | | | | | | | | | | | | | | |
| Unknown | 11 (100%) | 27% | 36% | 36% | 9% | 27% | 18% | 9% | 0% | 27% | 0% | 9% | 0% | 9% |
| 13-16 | 321 (100%) | 45% | 46% | 12% | 19% | 36% | 17% | 12% | 8% | 14% | 5% | 7% | 5% | 2% |
| 17-19 | 621 (100%) | 46% | 43% | 17% | 22% | 25% | 19% | 14% | 12% | 12% | 6% | 5% | 7% | 4% |
| 20-25 | 1468 (100%) | 48% | 37% | 27% | 23% | 15% | 15% | 14% | 15% | 9% | 7% | 5% | 4% | 6% |
| 25+ | 630 (100%) | 55% | 35% | 24% | 24% | 16% | 14% | 15% | 16% | 12% | 9% | 3% | 4% | 4% |
| **Respondent group** | | | | | | | | | | | | | | |
| Parent or expectant | 785 (100%) | 54% | 35% | 23% | 24% | 16% | 15% | 14% | 15% | 11% | 9% | 4% | 5% | 3% |
| Young person | 2,266 (100%) | 47% | 40% | 23% | 22% | 21% | 16% | 14% | 13% | 11% | 6% | 6% | 5% | 5% |

*Stratified percentages add up to over 100%

Supplementary Appendix 11: The percentage of total n respondents that reported better, worse, or no changes to their cities, stratified by $PM_{2.5}$ quartile, age bucket, and respondent group

| | Total n (%) | Becoming a nicer place to live | Staying the same | Becoming a worse place to live |
|---|---|---|---|---|
| **Full sample** | 2,993 (100%) | 43% | 23% | 34% |
| **$PM_{2.5}$ quartile** | | | | |
| 1 | 113 (100%) | 43% | 37% | 19% |
| 2 | 67 (100%) | 39% | 28% | 33% |
| 3 | 898 (100%) | 31% | 30% | 39% |
| 4 | 1,915 (100%) | 48% | 19% | 33% |
| **Age bucket** | | | | |
| Unknown | 11 (100%) | 18% | 45% | 36% |
| 13-16 | 312 (100%) | 45% | 29% | 26% |
| 17-19 | 611 (100%) | 43% | 26% | 31% |
| 20-25 | 1,437 (100%) | 44% | 22% | 34% |
| 25+ | 622 (100%) | 38% | 20% | 42% |
| **Respondent group** | | | | |
| Parent or expectant | 778 (100%) | 39% | 22% | 40% |
| Young person | 2,215 (100%) | 44% | 24% | 32% |

Supplementary Appendix 122: Mean air quality (scale 0-10) reported by n=3,054 respondents, stratified by $PM_{2.5}$ quartile

| | Total n (%) | Mean Air Quality (Likert Scale 0-10) | SD |
|---|---|---|---|
| **Full sample** | 3,054 (100%) | 5.07 | 2.86 |
| **$PM_{2.5}$ quartile** | | | |
| 1 | 113 (4%) | 6.01 | 2.19 |
| 2 | 67 (2%) | 5.31 | 2.42 |
| 3 | 897 (29%) | 5.73 | 2.64 |
| 4 | 1,977 (65%) | 4.71 | 2.94 |
| **Age bucket** | | | |
| Unknown | 11 (0.4%) | 4.73 | 3.07 |
| 13-16 | 320 (10%) | 5.25 | 2.81 |
| 17-19 | 622 (20%) | 4.97 | 2.88 |
| 20-25 | 1,469 (48%) | 5.11 | 2.89 |
| 25+ | 632 (21%) | 4.99 | 2.79 |
| **Respondent group** | | | |
| Parent or expectant | 787 (26%) | 5.04 | 2.76 |
| Young person | 2,267 (74%) | 5.08 | 2.9 |

Supplementary Appendix 13: The percentage of total n respondents who reported each major source of AP in their city, stratified by $PM_{2.5}$ quartile, age bucket, and respondent group

| | Total n (100%*) | Motor transport (cars, buses, lorries) | Factories | Burning of rubbish | Construction/building work | Pollution blown into the city (from outside) | Household cooking (cooking fires/stoves) | Household heating (boilers, wood fires etc) | Agriculture/farming | Other |
|---|---|---|---|---|---|---|---|---|---|---|
| **Full Sample** | 3,056 (100%) | 32% | 18% | 17% | 14% | 8% | 5% | 5% | 2% | 1% |
| **$PM_{2.5}$ Quartile** | | | | | | | | | | |
| 1 | 113 (100%) | 90% | 35% | 18% | 44% | 13% | 20% | 12% | 3% | 4% |
| 2 | 67 (100%) | 81% | 28% | 12% | 40% | 15% | 16% | 7% | 4% | 3% |
| 3 | 898 (100%) | 77% | 39% | 49% | 21% | 16% | 10% | 13% | 4% | 2% |
| 4 | 1,978 (100%) | 75% | 44% | 37% | 36% | 20% | 11% | 10% | 4% | 2% |
| **Age bucket** | | | | | | | | | | |
| Unknown | 11 (100%) | 64% | 45% | 55% | 18% | 9% | 0% | 9% | 9% | 9% |
| 13-16 | 321 (100%) | 79% | 38% | 36% | 33% | 18% | 12% | 6% | 2% | 2% |
| 17-19 | 623 (100%) | 74% | 50% | 39% | 34% | 19% | 11% | 9% | 4% | 1% |
| 20-25 | 1,470 (100%) | 74% | 42% | 42% | 32% | 19% | 13% | 12% | 4% | 2% |
| 25+ | 631 (100%) | 81% | 37% | 36% | 29% | 17% | 8% | 12% | 3% | 2% |
| **Respondent group** | | | | | | | | | | |
| Parent or expectant | 786 (100%) | 79% | 38% | 36% | 27% | 18% | 11% | 8% | 4% | 2% |
| Young person | 2,270 (100%) | 75% | 43% | 41% | 33% | 19% | 10% | 12% | 4% | 2% |

*Stratified percentages add up to over 100%

Supplemental Appendix 14: Illustrative quotes of youth's specific ideas and asks for their cities, by subtheme

**Concerns about inequality**
“Easy Living For Rich People, Hard for Poor or Middle Class Family”

**Corruption and bad governance**
“Corruption is the root of all these problems. None can solve these issues without solving corruption”

**Young people being absent from decision making**
“Engage the young people to be in the front"

Supplemental Appendix 15: Illustrative quotes of structural barriers to change, by subtheme

**City Design and Space**
*“There is no other way than planting more trees”*
*“Building playground, parks, and offering clean and safe environment”*

**Urban Mobility**
*“Creating a massive, comfortable, fast and efficient transportation system”*
*“Make more roads into pedestrian areas and build more bike lanes.”*

**Health**
*“Healthcare must be free of cost especially primary health care”*
*“Health care system is a mess, if you don’t have money you’ll die because they won’t attend to you”*

**Education**
*“…improve the education system by making it more fun to learn…”*
*“For young: quality education, proper knowledge of how contemporary world and near future world works.”*

**Skills and jobs**
*“We need job opportunities”*
*“I would firstly improve job opportunities and salary's cause one can only establish a stable life when they have a house as an asset and not liability and have a constant source of income which they use to establish or live an average life without the fear of poverty.”*

**Other basic services**
*“Improve housing, food and water supply”*
*“basic services such as clean water and or electricity are not consistent or available for a lot of people. There is no refuse collection and raw sewerage flowing in many areas.”*
